# Validity of European-centric derived cardiometabolic polygenic risk scores in multi-ancestry populations

**DOI:** 10.1101/2023.03.20.23287475

**Authors:** Constantin-Cristian Topriceanu, Nish Chaturvedi, Rohini Mathur, Victoria Garfield

**Author notes:** **Corresponding author** Dr. Constantin-Cristian TOPRICEANU, Research Fellow, UCL MRC Unit for Lifelong Health ang Aging, 1-19 Torrington Place, London, WC1E 7HB. Joint last authors (Rohini Mathur, Victoria Garfield).

## Abstract

Polygenic risk scores (PRSs) provide an individual level estimate of genetic risk for any given disease. Since most PRSs have been derived from genome wide association studies (GWAS) conducted in populations of White European ancestry, their validity in other ancestry groups remains unconfirmed. This is especially relevant for cardiometabolic diseases which are known to disproportionately affect people of non-European ancestry. Thus, we aimed to evaluate the performance of PRSs for glycaemic traits (glycated haemoglobin, type 1, and type 2 diabetes mellitus), cardiometabolic risk factors (body mass index, hypertension, high- and low-density lipoproteins, and total cholesterol and triglycerides) and cardiovascular diseases (CVDs, including stroke and coronary artery disease) in people of White European, South Asian, and African Caribbean ethnicity in the UK Biobank. Whilst the PRSs incorporated some GWAS data from multi-ethnic population, the vast majority originated from White Europeans. Except for hypertension and stroke, PRSs derived mostly from European populations had an overall better performance in White Europeans compared to South Asians and African Caribbeans. Thus, multi-ancestry GWAS data are needed to generate ancestry stratified PRSs to tackle health inequalities.

## INTRODUCTION

A polygenic risk score (PRS) provides a personalised estimate of an individual’s genetic liability to a disease. These are calculated as a weighted sum of single nucleotide polymorphisms (SNPs) ^1^. Although they are an exciting prospect for precision medicine, they might perpetuate health inequalities. Because most existing genome wide association studies (GWAS) studies have been conducted in populations of European ancestry^2,3^, their validity in other ancestry groups remains unconfirmed.

In genetic studies, ancestry is commonly used as a proxy for ethnicity. However, ethnicity is a complex concept which includes genetic ancestry and a wide range of social constructs (e.g., cultural practices, health beliefs, language, religion, and self-identification) ^4^. In general, ancestry is thought to better explain genetic relatedness and gene-environment interactions than ethnicity, due to the fact that ethnicity is a broader social concept which incorporates environmental measures of, socioeconomic status, lifestyle, etc^5^. However, there is a considerable overlap between ancestry and self-reported ethnicity, although ancestry does not capture the entirety of an individual’s ethnic identity.^6^ Thus, self-reported ethnicity is important when examining health disparities related to the wider socio-cultural and environmental determinants of health in addition to biological and genetic factors ^5^.

United States (US) data suggests that PRSs derived in European ancestry populations perform equally well in US Whites and Hispanic groups, but less well in African Americans^7^. Even then, within ethnicity heterogeneity can contribute to different predictive powers in ethnic sub-groups. For example, amongst Hispanics, the PRSs can have different performances based on ancestry clusters^8^. Thus, the transethnic transferability of PRSs remains a matter of debate.

Worldwide there are 500-700 million individuals with diabetes mellitus (DM), 90% of whom have type 2 diabetes (T2DM)^9^. The prevalence of T2DM differs by age (more common in older people), sex (more common in men) and ethnicity. In the United Kingdom, South Asians are more likely to suffer from diabetes ^10^, followed by those from an African Caribbean background ^11^ both of whom have 2-3 higher risk of developing T2DM compared to white groups ^11^. Individuals of South Asian and African Caribbean ethnicity are more likely to have higher serum glycated haemoblobin A1c [HbA1c] levels even in the absence of diabetes ^12^ and poorer glycaemic control in established diabetes^13^.

In addition to ethnic differences in diabetes risk, cardiometabolic risk factors and cardiovascular outcomes also vary across ethnicities. Compared to White Europeans, South Asians are more likely to develop cardiovascular disease (CVD; i.e., coronary artery disease [CAD] and stroke), whilst those from an African Caribbean background are more likely to suffer from strokes ^14^. Cardiovascular risk factors generally map to these ethnic differences in CVD outcomes. African Caribbeans generally have healthier lipid profiles (e.g., higher high-density lipoproteins [HDL] and lower total triglycerides [TTG] ^15^) compared to White Europeans and South Asians, in whom lipoprotein profiles are most adverse ^16^. In contrast, hypertension is more frequent in African Caribbeans than White Europeans ^17^. The picture is more complex for South Asians, who have an equivalent or lower blood pressure (BP) than Europeans at younger ages ^18^, but subsequently experience a steeper BP trajectory resulting in higher later life BP ^19^.

Whether PRSs derived mostly from white ancestry GWAS data can capture differences by self-reported ethnicity in glycaemic traits, cardiometabolic risk factors and cardiovascular outcomes remains unclear.

Using data from the UK Biobank (UKB), this study aimed to explore the prognostic value of transethnic transferability for a range of cardiometabolic PRSs and their respective observed outcomes. Our focus was on participants of South Asian and African Caribbean ethnicity in relation to White Europeans as these are the largest ethnic minority groups in the UK and are therefore well represented in UKB.

## METHODS

### Study population

The UKB is a large UK based prospective cohort study with >500000 participants recruited between 2006 and 2010 when study participants were aged 40-69 years old, and features demographic, genetic, health outcomes and imaging data for participants. ^20^. The breakdown of self-reported ethnicity in the UKB sample is 94.4% White Europeans, 0.2% South Asians, 0.2% African Caribbeans, and 5.2% other/unknown. Ancestry was previously derived in the UK Biobank using principal component analysis (PCA) and clustering, and it shows a good agreement with the self-reported ethnicity^21^.

### Data availability

The UK Biobank data is available via an application from https://www.ukbiobank.ac.uk/.

### Ethical approval

UK Biobank’s ethical approval (11/NW/0382) was from the Northwest Multi-centre Research Committee (MRCEC) in 2011, which was renewed in 2016 and then in 2021. All procedures performed were in accordance with the ethical standards of the institutional and/or national research committee and with the 1964 Helsinki declaration and its later amendments or comparable ethical standards.

### Exposure: Polygenic Risk Scores

Standard UK Biobank PRSs (n=36) include data from UK Biobank GWAS studies which contained 95% White individuals, while enhanced PRSs (n=53) contain in addition external GWAS data ^22^. In December 2022, we selected standard and enhanced PRSs related to glycaemic traits, associated cardiometabolic risk factors and cardiovascular outcomes namely: (1) T1DM, (2) T2DM, (3) glycated haemoglobin (HbA1c), (4) body mass index (BMI), (4) hypertension, (5) CVD, (6) CAD, (7) ischemic stroke, (8) HDL cholesterol, (9) low-density lipoprotein (LDL) cholesterol, (10) total cholesterol and (11) TTG. For total cholesterol and triglycerides, only an enhanced PRSs was available.

The methods employed to derive the PRSs have been previously described^22^. A custom Axion genotyping array (able to assay 825927 genetic variants) followed by genome-wide imputation was used to yield the genotype data. Ancestry classification was done using the same methodology which showed a good overlap between ethnicity and ancestry. The proportion of the genotype associated with White Europeans, South Asians and African Caribbeans ancestry was determined using the subset of common SNPs from the 1000 Genomes reference dataset, and genetic PCA was conducted to derive the centroid coordinates for ancestry groups, and hence to define the ancestry categories^21^. UK Biobank GWAS summary statistics were performed in PLINK 2.0 using logistic regression for binary categorical outcomes, and linear regression for continuous outcomes. When deriving the PRSs, the models adjusted for age, sex, genotyping chip, and ancestry principal components (PCs). In the study deriving the PRSs^22^, a literature review was employed to identify external GWAS summary statistics from the following studies: Atherosclerosis Risk in Communities (ARIC); Discovery, Biology and Risk of Inherited Variants in

Breast Cancer (DRIVE), Electronic Medical Records and Genomics (eMERGE), the Charles Bronfman Institute for Personalized Medicine (IPM) BioMe BioBank, Jackson Heart Study (JHS), Multi-Ethnic Cohort (MEC), the Multi-Ethnic Study of Atherosclerosis (MESA), Omics in Lations (OLA), GWAS for Breast Cancer in the African Diaspora (ROOT study). GWAS data were combined used a Bayesian fixed-effect inverse variance meta-analysis. UK Biobank and external GWAS data were meta-analyzed to yield the enhanced PRSs, whilst external GWAS data without UK Biobank data were combined to yield the standard PRSs. Lastly, a PC-based ancestry centering step was performed to zero center the PRSs across all ancestries^23^.

### Outcomes

All outcomes were evaluated using information captured at the baseline assessment between 2006-2010 in the 22 recruitment centres across England, Scotland, and Wales. Our primary outcomes were clinical phenotypes as recorded at baseline directly related to the PRSs. These included the presence of T1DM, T2DM, CVD, hypertension, and CAD (yes/no) as self-reported in health questionnaires; weight (kg), height (m), and blood pressure (mmHg) as recorded at the initial assessment, and HbA1c (mmol/mol), HDL (mmol/l), LDL (mmol/l), total cholesterol (mmol/l) and TTG (mmol/l) as taken from blood samples. The presence of hypertension at baseline was defined as either: (1) self-report of anti-hypertensive medication use, (2) systolic BP>140mmHg or (3) diastolic BP> 80mmHg. BMI (kg/m^2^) was calculated as the ratio of weight to height^2^.

### Covariates

Sex was self-reported as male or female, and age (years) was recorded at the time of recruitment. Area based Townsend deprivation scores were used to capture socio-economic position (SEP) ^24^. The primary care survey provided data on the prescribed medications of the study participants.

### Statistical Analysis

All analyses were performed in R 4.2.1 ^25^. Data distributions were assessed by histograms. Continuous variables were expressed as mean ± 1 standard deviation (SD) or median (interquartile range) as appropriate; categorical variables, as counts and percent.

Participants were categorized based on self-reported ethnicity as White European, South Asian, and African Caribbean. Individuals of mixed, other, and unknown ethnicity were not included due to small sample size. All analyses were conducted within each ethnic group. Generalised linear models (glms) with gamma distribution with identity link were used for continuous normally distributed outcomes. For binary outcomes, glms with binominal distribution (i.e., logistic regression) were employed.

Two regression models were compared. Model 1 was unadjusted to obtain raw estimates. As adjustment for genetic PCAs was previously used to control for ancestry during the PRS derivation process, further adjustment for principal components was not pursued. To obtain more precise regression estimates, Model 2 adjusted for factors associated with the outcome, namely age, sex, and SEP). For HbA1c, Model 2 additionally adjusted for diabetes medications. For HDL, LDL, total cholesterol and TTG, model 2 additionally adjusted for lipid-lowering medications. As those on antihypertensive medications were scored 1 for the hypertension outcome, model 2 for hypertension was not further adjusted for anti-hypertensive medication use. Since this study did not attempt to explore mechanistic pathways downstream of the genotype but upstream from the phenotypes, further models with adjustment for mediators were not pursued. Model assumptions were verified with regression diagnostics and found to be satisfied. Results were then corrected for multiple testing using a false discovery rate (FDR) of 0.05^26^.

We aimed to evaluate the performance of PRSs across all three ethnicities. For each binary outcome, we split each ethnicity specific dataset into 70% training and 30% testing. Using the training dataset, the logistic regression model was generated. Using the testing dataset, the classification performance (i.e., predicting the binary outcome) of the logistic regression model was evaluated using the receiver operating characteristic (ROC) curve. The area under the curve (AUC) and its associated 95% confidence interval (CI) was derived for each ethnicity for each binary outcome. The ROC curves were compared between ethnicities using DeLong’s test. For continuous outcomes, the exponentiated effect size captured the percentage difference in the outcome per unit increase in PRSs. Since the PRSs underwent a PC-based ancestry centering and had a normal distribution with a similar standard deviation (SD) (**Table 1**), the exponentiated effect size was not further standardized.

**Table 1.**
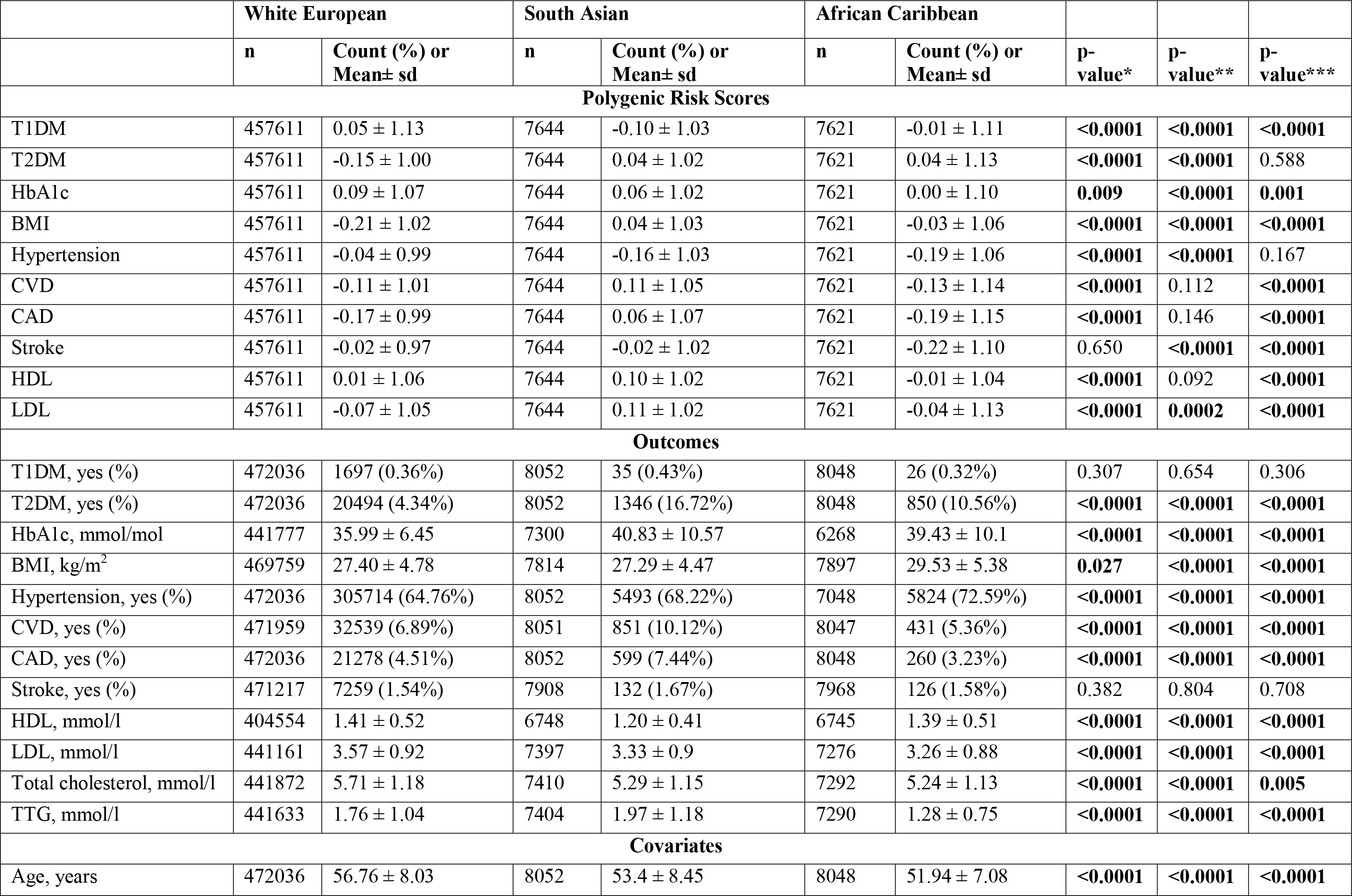
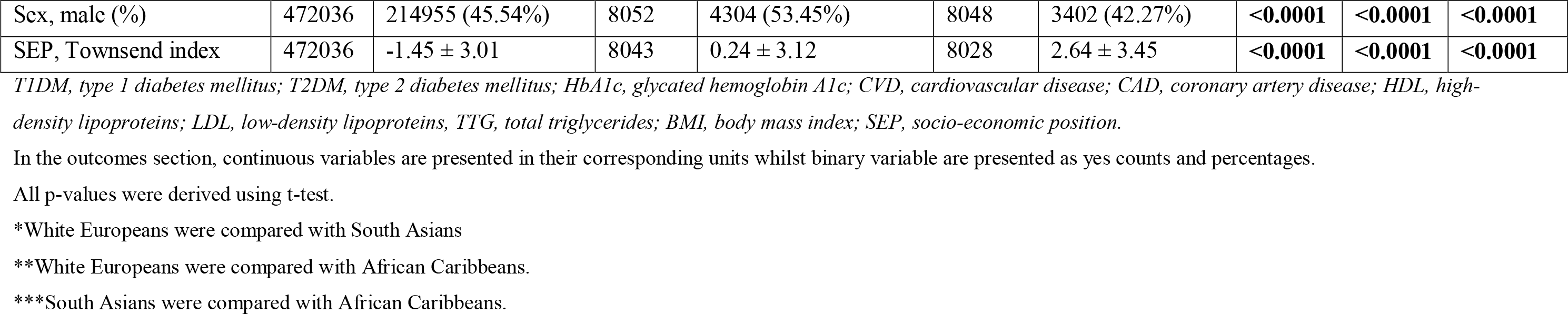
Participant characteristics per ethnic group for standard polygenic risk scores.

|  | White European |  | South Asian |  | African Caribbean |  |  |  |  |
| --- | --- | --- | --- | --- | --- | --- | --- | --- | --- |
|  | n | Count (%) or Mean± sd | n | Count (%) or Mean± sd | n | Count (%) or Mean± sd | p-value* | p-value** | p-value*** |
| **Polygenic Risk Scores** |  |  |  |  |  |  |  |  |  |
| T1DM | 457611 | 0.05 ± 1.13 | 7644 | -0.10 ± 1.03 | 7621 | -0.01 ± 1.11 | <0.0001 | <0.0001 | <0.0001 |
| T2DM | 457611 | -0.15 ± 1.00 | 7644 | 0.04 ± 1.02 | 7621 | 0.04 ± 1.13 | <0.0001 | <0.0001 | 0.588 |
| HbA1c | 457611 | 0.09 ± 1.07 | 7644 | 0.06 ± 1.02 | 7621 | 0.00 ± 1.10 | 0.009 | <0.0001 | 0.001 |
| BMI | 457611 | -0.21 ± 1.02 | 7644 | 0.04 ± 1.03 | 7621 | -0.03 ± 1.06 | <0.0001 | <0.0001 | <0.0001 |
| Hypertension | 457611 | -0.04 ± 0.99 | 7644 | -0.16 ± 1.03 | 7621 | -0.19 ± 1.06 | <0.0001 | <0.0001 | 0.167 |
| CVD | 457611 | -0.11 ± 1.01 | 7644 | 0.11 ± 1.05 | 7621 | -0.13 ± 1.14 | <0.0001 | 0.112 | <0.0001 |
| CAD | 457611 | -0.17 ± 0.99 | 7644 | 0.06 ± 1.07 | 7621 | -0.19 ± 1.15 | <0.0001 | 0.146 | <0.0001 |
| Stroke | 457611 | -0.02 ± 0.97 | 7644 | -0.02 ± 1.02 | 7621 | -0.22 ± 1.10 | 0.650 | <0.0001 | <0.0001 |
| HDL | 457611 | 0.01 ± 1.06 | 7644 | 0.10 ± 1.02 | 7621 | -0.01 ± 1.04 | <0.0001 | 0.092 | <0.0001 |
| LDL | 457611 | -0.07 ± 1.05 | 7644 | 0.11 ± 1.02 | 7621 | -0.04 ± 1.13 | <0.0001 | 0.0002 | <0.0001 |
| **Outcomes** |  |  |  |  |  |  |  |  |  |
| T1DM, yes (%) | 472036 | 1697 (0.36%) | 8052 | 35 (0.43%) | 8048 | 26 (0.32%) | 0.307 | 0.654 | 0.306 |
| T2DM, yes (%) | 472036 | 20494 (4.34%) | 8052 | 1346 (16.72%) | 8048 | 850 (10.56%) | <0.0001 | <0.0001 | <0.0001 |
| HbA1c, mmol/mol | 441777 | 35.99 ± 6.45 | 7300 | 40.83 ± 10.57 | 6268 | 39.43 ± 10.1 | <0.0001 | <0.0001 | <0.0001 |
| BMI, kg/m<sup>2</sup> | 469759 | 27.40 ± 4.78 | 7814 | 27.29 ± 4.47 | 7897 | 29.53 ± 5.38 | 0.027 | <0.0001 | <0.0001 |
| Hypertension, yes (%) | 472036 | 305714 (64.76%) | 8052 | 5493 (68.22%) | 7048 | 5824 (72.59%) | <0.0001 | <0.0001 | <0.0001 |
| CVD, yes (%) | 471959 | 32539 (6.89%) | 8051 | 851 (10.12%) | 8047 | 431 (5.36%) | <0.0001 | <0.0001 | <0.0001 |
| CAD, yes (%) | 472036 | 21278 (4.51%) | 8052 | 599 (7.44%) | 8048 | 260 (3.23%) | <0.0001 | <0.0001 | <0.0001 |
| Stroke, yes (%) | 471217 | 7259 (1.54%) | 7908 | 132 (1.67%) | 7968 | 126 (1.58%) | 0.382 | 0.804 | 0.708 |
| HDL, mmol/l | 404554 | 1.41 ± 0.52 | 6748 | 1.20 ± 0.41 | 6745 | 1.39 ± 0.51 | <0.0001 | <0.0001 | <0.0001 |
| LDL, mmol/l | 441161 | 3.57 ± 0.92 | 7397 | 3.33 ± 0.9 | 7276 | 3.26 ± 0.88 | <0.0001 | <0.0001 | <0.0001 |
| Total cholesterol, mmol/l | 441872 | 5.71 ± 1.18 | 7410 | 5.29 ± 1.15 | 7292 | 5.24 ± 1.13 | <0.0001 | <0.0001 | 0.005 |
| TTG, mmol/l | 441633 | 1.76 ± 1.04 | 7404 | 1.97 ± 1.18 | 7290 | 1.28 ± 0.75 | <0.0001 | <0.0001 | <0.0001 |
| **Covariates** |  |  |  |  |  |  |  |  |  |
| Age, years | 472036 | 56.76 ± 8.03 | 8052 | 53.4 ± 8.45 | 8048 | 51.94 ± 7.08 | <0.0001 | <0.0001 | <0.0001 |
| Sex, male (%) | 472036 | 214955 (45.54%) | 8052 | 4304 (53.45%) | 8048 | 3402 (42.27%) | **<0.0001** | **<0.0001** | **<0.0001** |
| SEP, Townsend index | 472036 | -1.45 ± 3.01 | 8043 | 0.24 ± 3.12 | 8028 | 2.64 ± 3.45 | **<0.0001** | **<0.0001** | **<0.0001** |

*T1DM, type 1 diabetes mellitus; T2DM, type 2 diabetes mellitus; HbA1c, glycated hemoglobin A1c; CVD, cardiovascular disease; CAD, coronary artery disease; HDL, high-density lipoproteins; LDL, low-density lipoproteins, TTG, total triglycerides; BMI, body mass index; SEP, socio-economic position.*
In the outcomes section, continuous variables are presented in their corresponding units whilst binary variable are presented as yes counts and percentages.
All p-values were derived using t-test.
\*White Europeans were compared with South Asians
\*\*White Europeans were compared with African Caribbeans.
\*\*\*South Asians were compared with African Caribbeans.

## RESULTS

Participant characteristics and standard PRSs by ethnicity are presented in **Table 1**, while enhanced PRSs are presented in **Supplementary Table S1**. On average, both South Asians (53.4 years) and African Caribbeans (51.9 years) were younger than White Europeans (56.8 years) at the time of outcome assessment. Men comprised 45.5% of White, 54.5% of South Asians and 42.3% of African Caribbeans in the sample. There were 45.7% South Asians, 70.5% African Caribbeans, and 23.2% White in the lowest quartile of the Townsend deprivation index. South Asians had the highest prevalence of T2DM (16.7%), CVD (10.1%), and CAD (7.4%), whilst African Caribbeans had the highest average BMI (29.5) and the highest prevalence of hypertension (72.6%). Despite the PC-based ancestry centering, the PRSs experienced residual deviation from zero in South Asians and African Caribbeans (**Table 1**). Model 1 and Model 2 results for the standard and enhanced PRSs are presented in **Table 2**. Results from Model 2 for the enhanced PRSs are presented below.

**Table 2.** Regression results stratified per ethnicity.

|  |  |  | White European |  |  | South Asian |  |  | African Caribbean |  |  |
| --- | --- | --- | --- | --- | --- | --- | --- | --- | --- | --- | --- |
| Outcome | PRS type | Model | n | $\beta$ or OR (95% CI) | p-value | n | $\beta$ or OR (95% CI) | p-value | n | $\beta$ or OR (95% CI) | p-value |
| T1DM | Standard | Model 1 | 457611 | 2.85 (2.75, 2.96) | **<0.0001** | 7644 | 1.49 (1.08, 2.01) | **0.013** | 7621 | 1.35 (0.96, 1.87) | 0.080 |
|  |  | Model 2 | 457072 | 2.87 (2.76, 2.98) | **<0.0001** | 7635 | 1.50 (1.09, 2.04) | **0.011** | 7604 | 1.36 (0.96, 1.89) | 0.764 |
|  | Enhanced | Model 1 | 76877 | 3.05 (2.78, 3.35) | **<0.0001** | 7637 | 1.51 (1.10, 2.05) | **0.009** | 7618 | 1.39 (0.99, 1.93) | 0.053 |
|  |  | Model 2 | 76793 | 3.09 (2.72, 3.40) | **<0.0001** | 7627 | 1.52 (1.11, 2.07) | **0.008** | 7601 | 1.40 (0.99, 1.95) | 0.520 |
| T2DM | Standard | Model 1 | 457611 | 2.34 (2.31, 2.38) | **<0.0001** | 7644 | 1.86 (1.74, 1.99) | **<0.0001** | 7621 | 1.47 (1.38, 1.58) | **<0.0001** |
|  |  | Model 2 | 457072 | 2.42 (2.38, 2.46) | **<0.0001** | 7635 | 2.00 (1.87, 2.15) | **<0.0001** | 7604 | 1.53 (1.43, 1.64) | **<0.0001** |
|  | Enhanced | Model 1 | 76877 | 2.23 (2.15, 2.31) | **<0.0001** | 7637 | 1.81 (1.70, 1.93) | **<0.0001** | 7601 | 1.38 (1.30, 1.48) | **<0.0001** |
|  |  | Model 2 | 76793 | 2.48 (2.39, 2.58) | **<0.0001** | 7628 | 2.05 (1.91, 2.20) | **<0.0001** | 7601 | 1.51 (1.41, 1.62) | **<0.0001** |
| HbA1c | Standard | Model 1 | 437520 | 0.84 (0.82, 0.85) | **<0.0001** | 7187 | 1.32 (1.07, 1.56) | **<0.0001** | 6214 | 0.48 (0.25, 0.72) | **<0.0001** |
|  |  | Model 2 | 436997 | 0.63 (0.62, 0.65) | **<0.0001** | 7179 | 0.82 (0.64, 0.99) | **<0.0001** | 6203 | 0.45 (0.28, 0.62) | **<0.0001** |
|  | Enhanced | Model 1 | 73455 | 1.69 (1.65, 1.73) | **<0.0001** | 7181 | 1.82 (1.60, 2.05) | **<0.0001** | 6211 | 1.07 (0.83, 1.30) | **<0.0001** |
|  |  | Model 2 | 73377 | 1.53 (1.50, 1.57) | **<0.0001** | 7173 | 1.30 (1.14, 1.46) | **<0.0001** | 6200 | 0.91 (0.75, 1.08) | **<0.0001** |
| BMI | Standard | Model 1 | 455985 | 1.32 (1.31, 1.33) | **<0.0001** | 7462 | 1.10 (1.00, 1.20) | **<0.0001** | 7505 | 0.73 (0.61, 0.85) | **<0.0001** |
|  |  | Model 2 | 455453 | 1.31 (1.30, 1.32) | **<0.0001** | 7453 | 1.08 (0.98, 1.18) | **<0.0001** | 7488 | 0.73 (0.62, 0.85) | **<0.0001** |
|  | Enhanced | Model 1 | 76572 | 1.72 (1.69, 1.74) | <**0.0001** | 7455 | 1.34 (1.24, 1.43) | <**0.0001** | 7502 | 0.91 (0.80, 1.02) | <**0.0001** |
|  |  | Model 2 | 76490 | 1.71 (1.68, 1.74) | <**0.0001** | 7446 | 1.31 (1.22, 1.40) | <**0.0001** | 7485 | 0.90 (0.80, 1.00) | <**0.0001** |
| Hypertension | Standard | Model 1 | 457611 | 1.52 (1.52, 1.54) | <**0.0001** | 7644 | 1.46 (1.38, 1.53) | <**0.0001** | 7621 | 1.19 (1.13, 1.25) | <**0.0001** |
|  |  | Model 2 | 457072 | 1.57 (1.56, 1.59) | <**0.0001** | 7635 | 1.51 (1.43, 1.60) | <**0.0001** | 7604 | 1.21 (1.15, 1.28) | <**0.0001** |
|  | Enhanced | Model 1 | 76877 | 1.63 (1.60, 1.65) | <**0.0001** | 7637 | 1.54 (1.47, 1.63) | <**0.0001** | 7618 | 1.28 (1.22, 1.34) | <**0.0001** |
|  |  | Model 2 | 76793 | 1.79 (1.76, 1.82) | <**0.0001** | 7628 | 1.67 (1.58, 1.77) | <**0.0001** | 7628 | 1.67 (1.58, 1.77) | <**0.0001** |
| CVD | Standard | Model 1 | 457539 | 1.49 (1.47, 1.51) | <**0.0001** | 7643 | 1.40 (1.30, 1.50) | <**0.0001** | 7620 | 1.13 (1.04, 1.24) | **0.0007** |
|  |  | Model 2 | 457001 | 1.53 (1.52, 1.55) | <**0.0001** | 7634 | 1.48 (1.37, 1.60) | <**0.0001** | 7603 | 1.14 (1.04, 1.25) | **0.007** |
|  | Enhanced | Model 1 | 76868 | 1.56 (1.51, 1.60) | <**0.0001** | 7636 | 1.47 (1.36, 1.59) | <**0.0001** | 7617 | 1.20 (1.09, 1.32) | **0.002** |
|  |  | Model 2 | 76784 | 1.61 (1.56, 1.66) | <**0.0001** | 7627 | 1.58 (1.45, 1.71) | <**0.0001** | 7600 | 1.20 (1.09, 1.32) | **0.0002** |
| CAD | Standard | Model 1 | 457611 | 1.71 (1.68, 1.73) | <**0.0001** | 7644 | 1.48 (1.36, 1.61) | <**0.0001** | 7621 | 1.18 (1.05, 1.32) | **0.004** |
|  |  | Model 2 | 457072 | 1.78 (1.75, 1.81) | <**0.0001** | 7635 | 1.55 (1.42, 1.69) | <**0.0001** | 7604 | 1.18 (1.06, 1.33) | **0.004** |
|  | Enhanced | Model 1 | 76877 | 1.83 (1.76, 1.90) | <**0.0001** | 7637 | 1.60 (1.47, 1.74) | <**0.0001** | 7618 | 1.21 (1.08, 1.36) | **0.001** |
|  |  | Model 2 | 76793 | 1.91 (1.84, 1.99) | <**0.0001** | 7628 | 1.70 (1.55, 1.86) | <**0.0001** | 7601 | 1.22 (1.08, 1.37) | **0.001** |
| Stroke | Standard | Model 1 | 456848 | 1.34 (1.31, 1.38) | <**0.0001** | 7514 | 1.34 (1.12, 1.61) | **0.001** | 7545 | 1.13 (0.95, 1.35) | 0.164 |
|  |  | Model 2 | 456310 | 1.34 (1.30, 1.37) | <**0.0001** | 7505 | 1.36 (1.14, 1.64) | **0.0008** | 7528 | 1.13 (0.95, 1.35) | 0.167 |
|  | Enhanced | Model 1 | 76703 | 1.43 (1.34, 1.52) | <0.0001 | 7507 | 1.39 (1.15, 1.67) | 0.0006 | 7542 | 1.23 (1.05, 1.45) | 0.012 |
|  |  | Model 2 | 76619 | 1.42 (1.33, 1.51) | <0.0001 | 7498 | 1.42 (1.17, 1.71) | 0.0003 | 7525 | 1.23 (1.04, 1.45) | 0.015 |
| HDL | Standard | Model 1 | 399315 | 0.14 (0.14, 0.14) | <0.0001 | 6607 | 0.11 (0.10, 0.12) | <0.0001 | 6610 | 0.10 (0.09, 0.11) | <0.0001 |
|  |  | Model 2 | 396414 | 0.13 (0.13, 0.13) | <0.0001 | 6373 | 0.11 (0.10, 0.12) | <0.0001 | 6431 | 0.10 (0.09, 0.11) | <0.0001 |
|  | Enhanced | Model 1 | 67042 | 0.14 (0.14, 0.14) | <0.0001 | 6602 | 0.11 (0.10, 0.12) | <0.0001 | 6607 | 0.09 (0.08, 0.10) | <0.0001 |
|  |  | Model 2 | 66465 | 0.13 (0.13, 0.14) | <0.0001 | 6368 | 0.11 (0.10, 0.11) | <0.0001 | 6428 | 0.09 (0.08, 0.10) | <0.0001 |
| LDL | Standard | Model 1 | 435509 | 0.24 (0.24, 0.24) | <0.0001 | 7240 | 0.16 (0.14, 0.18) | <0.0001 | 7135 | 0.15 (0.14, 0.17) | <0.0001 |
|  |  | Model 2 | 432326 | 0.27 (0.27, 0.28) | <0.0001 | 6988 | 0.19 (0.17, 0.20) | <0.0001 | 6940 | 0.17 (0.15, 0.19) | <0.0001 |
|  | Enhanced | Model 1 | 73124 | 0.27 (0.26, 0.27) | <0.0001 | 7234 | 0.18 (0.16, 0.20) | <0.0001 | 7132 | 0.22 (0.20, 0.23) | <0.0001 |
|  |  | Model 2 | 72506 | 0.30 (0.30, 0.31) | <0.0001 | 6982 | 0.22 (0.20, 0.23) | <0.0001 | 6937 | 0.24 (0.22, 0.26) | <0.0001 |
| Total cholesterol | Enhanced | Model 1 | 73261 | 0.28 (0.27, 0.29) | <0.0001 | 7248 | 0.19 (0.17, 0.22) | <0.0001 | 7146 | 0.20 (0.18, 0.23) | <0.0001 |
|  |  | Model 2 | 72640 | 0.31 (0.30, 0.32) | <0.0001 | 6996 | 0.22 (0.19, 0.24) | <0.0001 | 6951 | 0.22 (0.20, 0.24) | <0.0001 |
| TTG | Enhanced | Model 1 | 73203 | 0.24 (0.23, 0.25) | <0.0001 | 7243 | 0.28 (0.26, 0.30) | <0.0001 | 7144 | 0.09 (0.07, 0.11) | <0.0001 |
|  |  | Model 2 | 72583 | 0.23 (0.22, 0.23) | <0.0001 | 6992 | 0.28 (0.25, 0.30) | <0.0001 | 6949 | 0.09 (0.07, 0.10) | <0.0001 |

Model 1 was unadjusted to obtain raw estimates as the relationship between genotype and phenotype should be unconfounded.
Model 2 was adjusted for age, sex, and SEP to obtain more precise regression estimates. Model 2 for HbA1c was in addition adjusted for diabetes medications, while HDL, LDL, total cholesterol and TTG models were adjusted for lipid-lowering medications. As those on blood lowering medications were scored 1 for the hypertension outcome, the hypertension model 2 was not further adjusted for anti-hypertensives
$\beta$ , regression coefficient; CI, confidence interval; OR, odds ratio. Other abbreviations as in **Table 1**.

### Type 1 diabetes

The association between the enhanced PRSs and T1DM was strongest for White Europeans (OR 3.09 95% CI [2.72, 3.40]) followed by South Asians (OR 1.52 95% CI [1.11, 2.07]) and African Caribbeans (OR 1.40 95% CI [0.99, 1.95]) (**Figure 1A**). The PRS’ predictive performance was highest in White Europeans (AUC 82.85 95% CI [80.82,84.98]) followed by South Asians (AUC 66.84 95% CI [52.75,80.94) and African Caribbeans (AUC 58.10 95% CI [36.06, 80.14]) (**Table 3**).

**Table 3.**
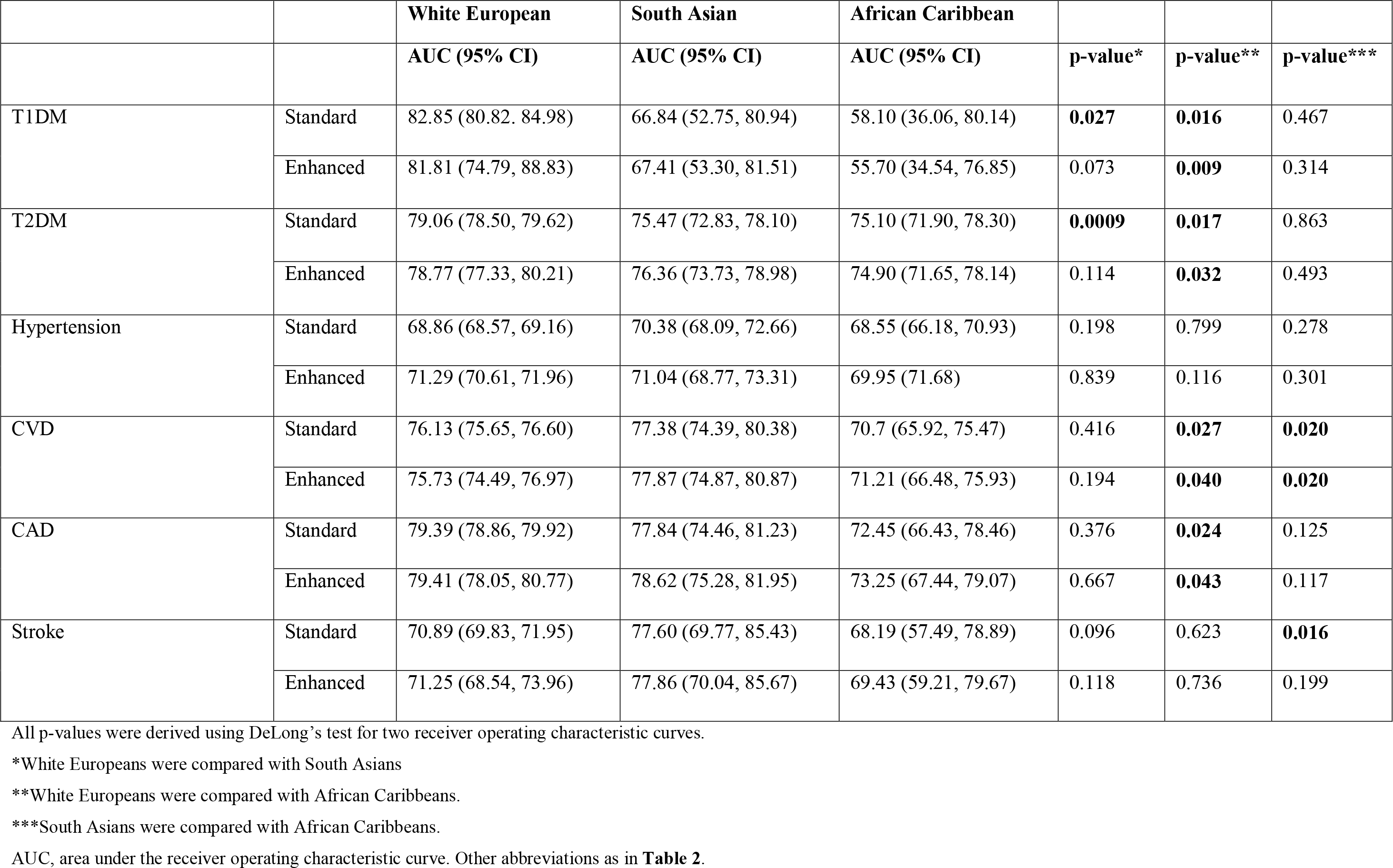
Predictive power of PRSs for diabetes-related binary outcomes stratified by ancestry in model 2.

|  |  | **White European** | **South Asian** | **African Caribbean** |  |  |  |
| --- | --- | --- | --- | --- | --- | --- | --- |
|  |  | **AUC (95% CI)** | **AUC (95% CI)** | **AUC (95% CI)** | **p-value\*** | **p-value\*\*** | **p-value\*\*\*** |
| T1DM | Standard | 82.85 (80.82, 84.98) | 66.84 (52.75, 80.94) | 58.10 (36.06, 80.14) | **0.027** | **0.016** | 0.467 |
|  | Enhanced | 81.81 (74.79, 88.83) | 67.41 (53.30, 81.51) | 55.70 (34.54, 76.85) | 0.073 | **0.009** | 0.314 |
| T2DM | Standard | 79.06 (78.50, 79.62) | 75.47 (72.83, 78.10) | 75.10 (71.90, 78.30) | **0.0009** | **0.017** | 0.863 |
|  | Enhanced | 78.77 (77.33, 80.21) | 76.36 (73.73, 78.98) | 74.90 (71.65, 78.14) | 0.114 | **0.032** | 0.493 |
| Hypertension | Standard | 68.86 (68.57, 69.16) | 70.38 (68.09, 72.66) | 68.55 (66.18, 70.93) | 0.198 | 0.799 | 0.278 |
|  | Enhanced | 71.29 (70.61, 71.96) | 71.04 (68.77, 73.31) | 69.95 (71.68) | 0.839 | 0.116 | 0.301 |
| CVD | Standard | 76.13 (75.65, 76.60) | 77.38 (74.39, 80.38) | 70.7 (65.92, 75.47) | 0.416 | **0.027** | **0.020** |
|  | Enhanced | 75.73 (74.49, 76.97) | 77.87 (74.87, 80.87) | 71.21 (66.48, 75.93) | 0.194 | **0.040** | **0.020** |
| CAD | Standard | 79.39 (78.86, 79.92) | 77.84 (74.46, 81.23) | 72.45 (66.43, 78.46) | 0.376 | **0.024** | 0.125 |
|  | Enhanced | 79.41 (78.05, 80.77) | 78.62 (75.28, 81.95) | 73.25 (67.44, 79.07) | 0.667 | **0.043** | 0.117 |
| Stroke | Standard | 70.89 (69.83, 71.95) | 77.60 (69.77, 85.43) | 68.19 (57.49, 78.89) | 0.096 | 0.623 | **0.016** |
|  | Enhanced | 71.25 (68.54, 73.96) | 77.86 (70.04, 85.67) | 69.43 (59.21, 79.67) | 0.118 | 0.736 | 0.199 |

All p-values were derived using DeLong's test for two receiver operating characteristic curves.
\*White Europeans were compared with South Asians
\*\*White Europeans were compared with African Caribbeans.
\*\*\*South Asians were compared with African Caribbeans.
AUC, area under the receiver operating characteristic curve. Other abbreviations as in **Table 2**.

**Figure 1.**
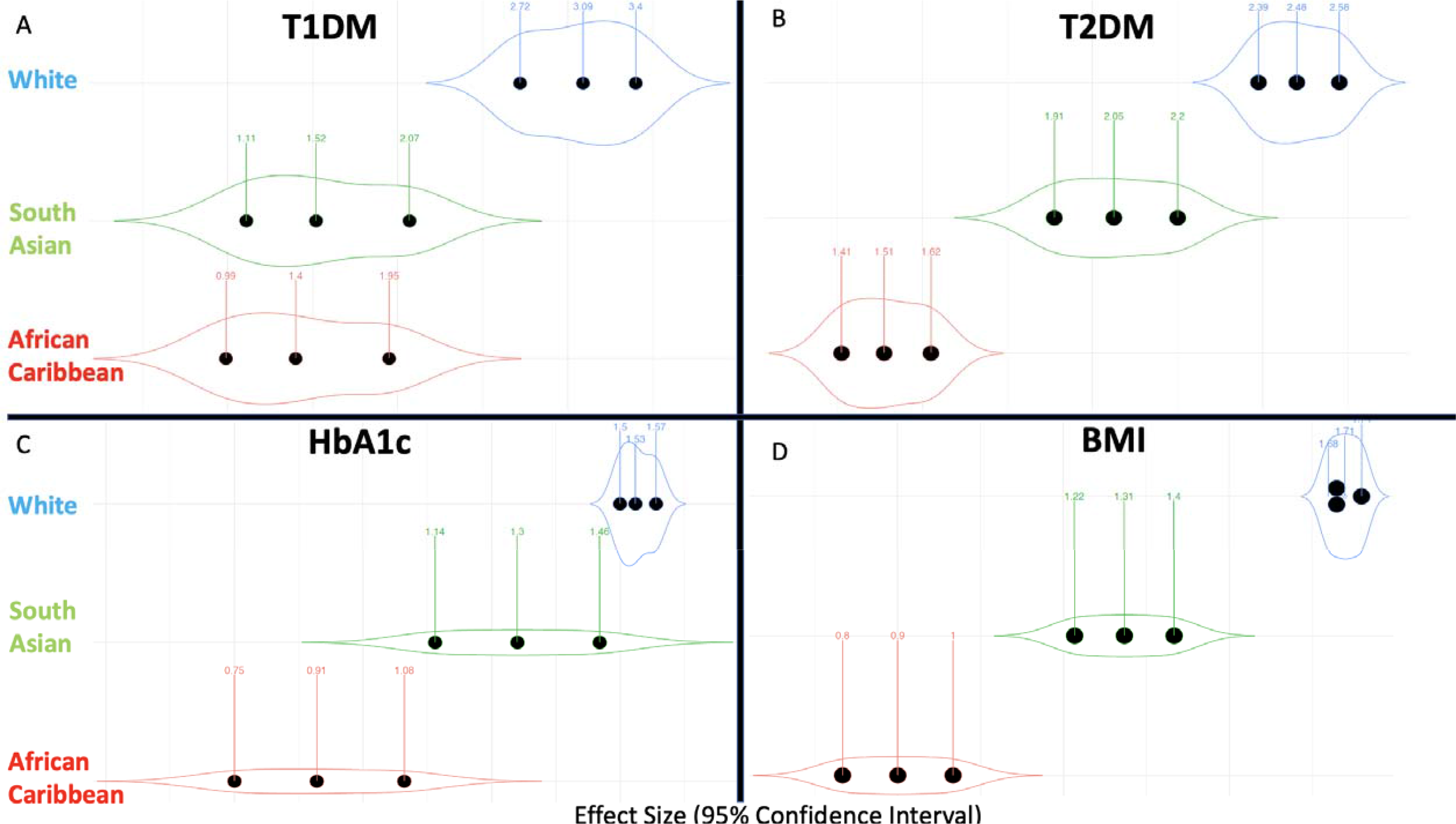
Violin plots highlighting the effect sizes per standard deviation increase in the enhanced PRS in model 2 for diabetes and body mass index by ethnicity. In each violin plot, the middle dot represents the effect size, and the adjacent ones the lower and upper limits of the 95% confidence intervals. The violin shape reflects the wideness of the confidence interval. *BMI, body mass index; HbA1c, glycosylated haemoglobinA1c; T1DM, type 1 diabetes mellitus; T2DM, type 2 diabetes mellitus*

### Type 2 diabetes

According to the OR, the highest performance was highest in White Europeans (OR 2.48 95% CI [2.39, 2.58]) followed by South Asians (OR 2.05 95% CI [1.91, 2.20]) and African Caribbeans (OR 1.51 95% CI 1.51 [1.30, 1.48]) (**Figure 1B**). According to the AUC, the enhanced PRS’ predictive performance was higher in White Europeans (AUC 78.77 95% CI [77.33, 80.21]) compared to African Caribbeans (AUC 74.90 95% CI [71.65, 78.14]) (**Table 3**).

### HbA1c

The regression coefficient (β) was highest in White Europeans followed by South Asians and African Caribbeans. One unit (or 1 SD) increase in the enhanced PRS was associated with a 1.53 mmol/mol 95% CI (1.50, 1.57) higher HbA1c in White Europeans, 1.30 95% CI (1.14, 1.46) in South Asians and 0.91 (0.75, 1.08) in African Caribbeans after adjusting for sex, age, SEP, and the presence of diabetes medications. Results are visually depicted in **Figure 1C**.

### BMI

A unit increase in the enhanced PRS resulted in a 1.71 kg/m^2^ difference 95% CI (1.68, 1.74) in BMI in White Europeans, 1.31 95% CI (1.22, 1.40) in South Asians and 0.90 95% CI (0.80, 1.00) in African Caribbeans (**Table 2 and Figure 1D**).

### CVD and CAD

For CVD, the performance of the enhanced PRS was higher in White Europeans (AUC 75.73 95% CI [74.49, 76.97]) and South Asians (AUC 77.87 95% CI [74.87, 80.87]) vs African Caribbeans (AUC 71.21 95% CI [66.48, 75.93]) all p<0.040 (**Table 3**). Similarly, the ORs were higher in White Europeans (OR 1.61 95% CI [1.56, 1.66]) and South Asians (OR 1.58 95% CI [1.45, 1.71]) compared to African Caribbeans (OR 1.20 95% CI [1.09, 1.32]) (**Figure 2A**).

**Figure 2.**
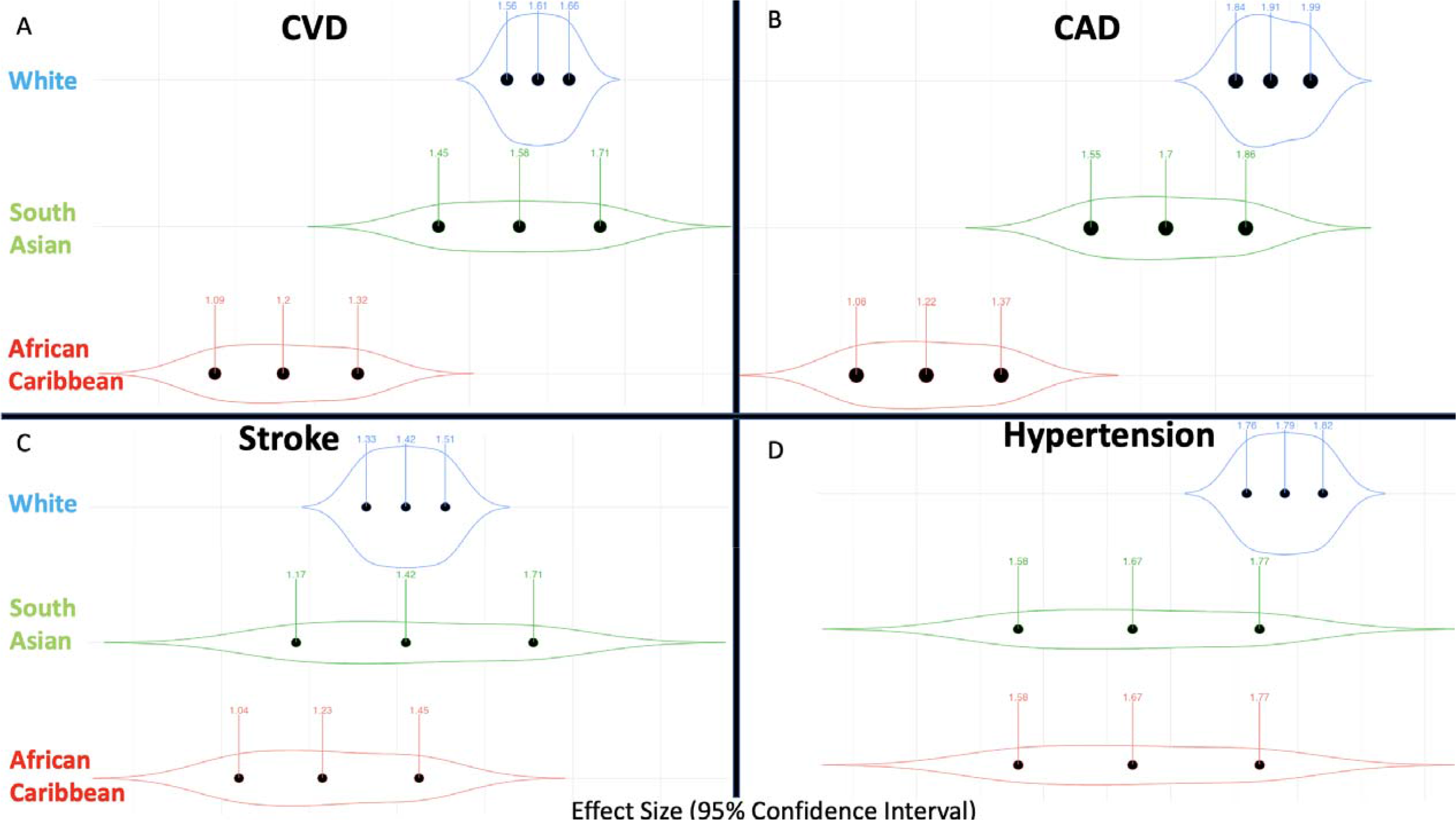
Violin plots highlighting the effect sizes per standard deviation increase in the enhanced PRS in model 2 for vessel-related outcomes. In each violin plot, the middle dot represents the effect size, and the adjacent ones the lower and upper limits of the 95% confidence intervals. The violin shape reflects the wideness of the confidence interval. *CAD, coronary artery disease; CVD, cardiovascular disease*.

For CAD, the results were similar to those reported above for CVD, with a higher predictive performance according to the ROC curve analysis (**Table 3**) and higher OR (**Table 2**) in White Europeans and South Asians compared to African Caribbeans.

### Hypertension

There was no difference in performance by ethnicity according to the ROC curve analysis (**Table 3**). The ORs were similar across all ethnicities using both standard (≈1.50) and enhanced PRSs (≈1.70). (**Table 2** and **Figure 2C**).

### Stroke

The performance of the enhanced PRS according to the ROC curve analysis (AUC ≈70) and the ORs (1.20-1.40) were similar across all ethnicities (**Figure 2D, Table 3**).

### HDL and LDL

One SD increase in the enhanced HDL PRS resulted in a 0.13 95% CI (0.13,0.14) greater HDL in White Europeans, 0.11 95% CI (0.10, 0.11) in South Asians and 0.09 95% CI (0.08, 0.10) in African Caribbeans (**Table 2 and Figure 3 A/B**) after adjusting for sex, age, SEP, and the presence of lipid-lowering drugs. A unit increase in the enhanced LDL PRS was associated with a higher LDL in White Europeans (0.30 95% CI [0.30, 0.31]) followed by African Caribbeans (0.24 95% CI [0.22, 0.26]) and South Asians (0.22 95% CI [0.20, 0.23]).

**Figure 3.**
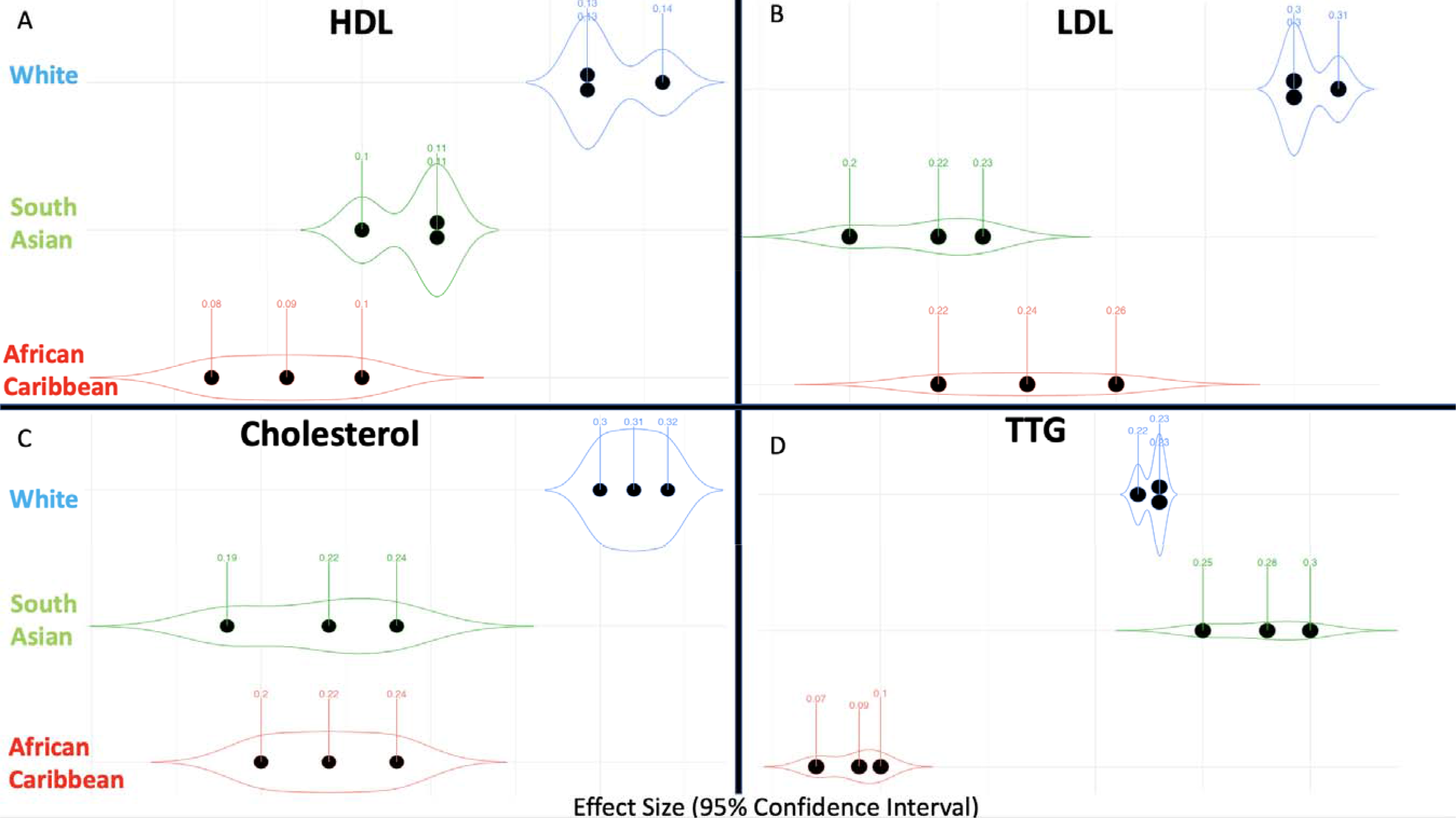
Violin plots highlighting the effect sizes per standard deviation increase in the enhanced PRS in model 2 for lipid-related outcomes. In each violin plot, the middle dot represents the effect size, and the adjacent ones the lower and upper limits of the 95% confidence intervals. The violin shape reflects the wideness of the confidence interval. *HDL, high-density lipoproteins; LDL, low-density lipoproteins; TTG, total triglycerides*.

### Total cholesterol and triglycerides

A unit (or 1 SD) increase in the enhanced PRS resulted in a greater total cholesterol in White Europeans (β≈0.30) compared to South Asians and African Caribbeans (β≈0.20) (**Figure 3C**). On the other hand, a unit (or 1SD) increase in the TTG enhanced PRS was associated with a higher TTG in White Europeans and South Asians (β≈0.25) compared to African Caribbeans (β≈0.10) (**Figure 3D**).

In this study we evaluated the performance of standard and enhanced PRSs for glycaemic traits, associated cardiometabolic risk factors and cardiovascular outcomes derived mostly in White European populations in association with their respective observed phenotype by ethnicity in the UKB. Whilst the UK Biobank PRSs included some data from multi-ethnic GWAS studies, the performance of both the standard and enhanced PRSs was better in those of White European ethnicity compared to those of South Asians and African Caribbean ethnicity for most outcomes except stroke and hypertension. This can be explained by the predominance of White European GWAS data when deriving the PRSs.

### Factors driving poorer PRS performance in ethnic diverse populations

According to the National Human Genome Research Institute (NHGRI) and European Bioinformatics Institute (EBI) GWAS catalogue almost 80% of the GWAS studies have been performed in White Europeans even though 25% of the global population is of South Asian ancestry and 15% is of African Caribbean ancestry^27^. Thus, the GWAS data is scarce in non-White ancestries. This has multiple downstream implications and might partly explain the worse performance in PRSs in multi-ethnic populations^28^. Firstly, linkage disequilibrium (LD) varies across ancestries meaning that these differences may drive disparities between effect size estimates in GWASs^29^. As PRSs are weighted sums, these errors are additively cumulated across ancestries which might explain the poorer performance in ethnic minorities ^2^. Secondly, imputation reference panels widely used to address bias in GWASs are less efficient in non-White ancestries due to data scarcity. Thirdly, within-ethnicity heterogeneity leading to differential predictive power in the same ethnicity has been reported ^8^, and within-ethnicity ancestry subcategories are even less studied. Finally, the normal reference ranges for quantitative biomarkers may vary between ethnicities^30^. Without ethnicity specific reference ranges and cut-offs, there is an inherent bias in any GWAS which categorizes/binarizes quantitative traits. Lastly, studies may be reporting common benign variants in other ethnicities as being pathologic because they are rare in white participants^2^. Thus, large ethnic diverse datasets and improved treatment of LD and variant frequencies are increasingly needed to create equitable PRSs before widespread clinical use^28^.

### Ethnic inclusivity for equitable implementation of polygenic risk scores

In CVD research, the vast majority of cohort studies enrolled mostly people from White European ancestry. There are only a few studies which include genetic data in ethnic minorities which can contain a single ethnic group (e.g., Genes & Health, China Kadoorie Biobank [CKB], Mexico City Prospective Study [MCPS], New Delhi Birth Cohort Study, OLA etc.) or multiple ethnic groups (e.g., Age, Gene/Environment Susceptibility [AGES], ARIC, Born in Bradford (BiB), Cardiovascular Health Study [CHS], Dallas Heart Study [DHS], Framingham Heart Study [FHS] OMNI cohorts, JHS, MEC, MESA, Rotterdam Study [RS], Southall and Brent Revisited [SABRE] etc.). Moreover, there is a tendency to aggregate individual cohorts into consortia (e.g., genetic data from AGES, ARIC, CHS, FHS and RS cohorts are available through the Cohorts for Heart and Aging Research in Genomic Epidemiology [CHARGE] consortium). Despite these collections, the percentage of non-White European ancestry participants in GWASs has not increased in recent years ^27^. This suggests that the reduced performance of PRSs in ethnic minorities is unlikely to improve in the near future.

The low participation by ethnic minorities in biomedical research is multi-factorial but mainly related to reduced trust given past research misconduct and feelings of racial discrimination ^31^. However, movements such as the All of Us research programme from National Institute of Health are working towards having a culturally aware approach to engage under-represented ethnic minorities in research^32^.

### Ancestry inclusivity for equitable implementation of polygenic risk scores

Race and ethnicity are socio-cultural constructs, whilst ancestry refers to the genetic origin of a population. ^4^Engaging under-represented ethnic and ancestry minorities in genomics research should be a global research priority. Indeed, there are movements aiming to address these disparities such as the Human Heredity and Health in Africa initiative ^33^. However, lack of funding remains the main limitation of international movements ^34^.

### Polygenic risk scores and health inequalities during translation to practice

The advent of genetic data in large cohort datasets such as the UK Biobank has led to the discovery of multiple SNPs which are associated with a variety of diabetes and cardiovascular related outcomes in GWAS. Whilst the added value of PRSs on top of already validated clinical tools is yet to be fully elucidated, current studies suggest that PRSs could: (1) increase disease prediction in early life, (2) help guide population-wide screening and preventative targeted interventions (e.g., lipid lowering drugs with those with a high PRS for total cholesterol and LDL), (3) help promote favourable health behaviours in those with an enhanced risk, (4) improve the diagnostic accuracy (e.g., differentiating T1DM vs T2DM in overweight antibody-negative young individuals), and (5) predicting response to treatments^35^. Given the worse performance of PRSs in ethnic minorities, they may miss out on benefiting from improved health outcomes. The deployment of PRSs would benefit the population group which is already privileged in terms of health outcomes further deepening existing healthcare inequalities. Thus, multi-ancestry GWAS data are needed to generate ethnicity stratified PRSs to tackle health inequalities.

### Limitations

Limitations of the UK Biobank PRSs have been previously discussed ^22^. Briefly, these limitations include: (1) the inclusion of GWAS data from predominantly White Europeans, (2) differences in how outcomes were defined between the studies included, and (3) cross cohort heterogeneity of PRSs. With regards to the PRS evaluation, the main limitation of our study relates to the lack of widely accepted performance metrics ^36^. Whilst the phenotypic variance explained (R^2^) and the association p-values have been proposed ^37^, we used effect-size metrics for the outcome as these are widely used for established traditional risk factors. However, these do not accurately capture disease prevalence in the general population.

## CONCLUSION

In general, UK Biobank standard and enhanced PRSs had markedly better performance in White Europeans compared to South Asians and African Caribbeans when evaluating cardiometabolic phenotypes. More GWAS data in ethnic minorities is required to improve the performance of the PRSs to avoid perpetuating health inequalities especially since diabetes is more prevalent in South Asians and African Caribbeans.

## Data Availability

The UK Biobank data is available via an application from https://www.ukbiobank.ac.uk/.

## DECLARATIONS

## CONFLICT OF INTEREST DISCLOSURES

The views expressed in this article are those of the authors who declare that they have no conflict of interest except for Nish Chaturvedi who serves on a Data Safety and Monitoring Board for a clinical trial of a glucose lowering agent, funded by AstraZeneca.

## FUNDING

NC received support from the UK Medical Research Council, Diabetes UK, Wellcome Trust, British Heart Foundation and National Institute for Health Research University College London Hospitals Biomedical Research Centre. RM is supported by Barts Charity (MGU0504). VG is funded by the Professor David Matthews Non-Clinical Fellowship (ref: SCA/01/NCF/22). VG is also supported by joint Diabetes UK and British Heart Foundation grant (ref: 15/0005250).

## ROLE OF THE FUNDING SOURCE

None of the funders was involved in the study design, the collection, the analysis, the interpretation of the data, and in the decision to submit the article for publication.

For the purpose of open access, the authors have applied a creative commons attribution (CC BY) license to any author accepted manuscript version arising.

## CONTRIBUTORS’ STATEMENT

All authors were involved in study design and implementation, data analysis and interpretation, critically reviewing and revising the manuscript. In addition, all authors approved the final version as submitted and agree to be accountable for all aspects of the work.

## ACKNOWLEDGEMENTS

The authors would like to thank all the UK Biobank members for their participation and continuous engagement with follow-up and all UK Biobank scientific and data collection teams.

## SUPPLEMENTAL MATERIALS

**Supplementary Table S1**

