## Supplementary Table S1 for "Validity of European-centric derived cardiometabolic polygenic risk scores in multi-ancestry populations"

*Joint last authors

**Author Affiliations:**

1. Department of Population Science and Experimental Medicine, Institute of Cardiovascular Science, University College London, Gower Street, London, WC1E 6BT, UK.
2. MRC Unit for Lifelong Health and Ageing, University College London, 1-19 Torrington Place, London, WC1E 7HB
3. Centre for Primary Care, Wolfson Institute of Population Health, Queen Mary University of London

**Corresponding author**

Dr. Constantin-Cristian TOPRICEANU

Research Fellow, UCL MRC Unit for Lifelong Health ang Aging

1-19 Torrington Place, London, WC1E 7HB

**Supplementary Table S1. Enhanced polygenic risk scores per ethnicity in UK Biobank.**

|  | **White European** | | **South Asian** | | **African Caribbean** | |  |  |  |
| --- | --- | --- | --- | --- | --- | --- | --- | --- | --- |
|  | **n** | **Count (%) or Mean± sd** | **n** | **Count (%) or Mean± sd** | **n** | **Count (%) or Mean± sd** | **p-value*** | **p-value**** | **p-value***** |
| **Polygenic Risk Scores** | | | | | | | | | |
| T1DM | 76877 | 0.00 ± 1.11 | 7637 | -0.09 ± 1.04 | 7618 | 0.00 ± 1.11 | **<0.0001** | 0.883 | **<0.0001** |
| T2DM | 76877 | -0.12 ± 1.04 | 7637 | 0.03 ± 1.02 | 7618 | 0.04 ± 1.16 | **<0.0001** | **<0.0001** | 0.351 |
| HbA1c | 76877 | -0.05 ± 1.09 | 7637 | 0.11± 1.10 | 7618 | -0.11 ± 1.11 | **<0.0001** | **<0.0001** | **<0.0001** |
| BMI | 76877 | -0.01 ± 1.06 | 7637 | -0.09 ± 1.06 | 7618 | -0.07 ± 1.14 | **<0.0001** | **<0.0001** | 0.231 |
| Hypertension | 76877 | -0.02 ± 1.01 | 7637 | -0.17 ± 1.02 | 7618 | -0.13 ± 1.11 | **<0.0001** | **<0.0001** | **0.017** |
| CVD | 76877 | -0.08 ± 1.04 | 7637 | 0.05 ± 1.03 | 7618 | -0.17 ± 1.12 | **<0.0001** | **<0.0001** | **<0.0001** |
| CAD | 76877 | -0.12 ± 1.02 | 7637 | -0.02 ± 1.07 | 7618 | -0.18 ± 1.13 | **<0.0001** | **<0.0001** | **<0.0001** |
| Stroke | 76877 | 0.01 ± 0.99 | 7637 | -0.09 ± 1.00 | 7618 | -0.22 ± 1.16 | **<0.0001** | **<0.0001** | **<0.0001** |
| HDL | 76877 | -0.02 ± 1.07 | 7637 | 0.16 ± 1.06 | 7618 | 0.01 ± 1.08 | **<0.0001** | **0.042** | **<0.0001** |
| LDL | 76877 | -0.01 ± 1.08 | 7637 | 0.19 ± 1.03 | 7618 | -0.09 ± 1.09 | **<0.0001** | **<0.0001** | **<0.0001** |
| Total cholesterol | 76877 | -0.07 ± 1.03 | 7637 | 0.17 ± 1.01 | 7618 | -0.07 ± 1.08 | **<0.0001** | 0.760 | **<0.0001** |
| TTG | 76877 | -0.07 ± 1.03 | 7644 | 0.17 ± 1.01 | 7618 | -0.07 ± 1.07 | **<0.0001** | 0.760 | **<0.0001** |

*T1DM, type 1 diabetes mellitus; T2DM, type 2 diabetes mellitus; HbA1c, glycated hemoglobin A1c; CVD, cardiovascular disease; CAD, coronary artery disease; HDL, high-density lipoproteins; LDL, low-density lipoproteins, TTG, total triglycerides; BMI, body mass index; SEP, socio-economic position.*

All p-values were derived using t-test.

*White Europeans were compared with South Asians

**White Europeans were compared with African Caribbeans.

***South Asians were compared with African Caribbeans.
